# Guidelines For Rigorous Evaluation of Clinical LLMs For Conversational Reasoning

**DOI:** 10.1101/2023.09.12.23295399

**Authors:** Shreya Johri, Jaehwan Jeong, Benjamin A. Tran, Daniel I. Schlessinger, Shannon Wongvibulsin, Zhuo Ran Cai, Roxana Daneshjou, Pranav Rajpurkar

## Abstract

The integration of Large Language Models (LLMs) like GPT-4 and GPT-3.5 into clinical diagnostics has the potential to transform patient-doctor interactions. However, the readiness of these models for real-world clinical application remains inadequately tested. This paper introduces the Conversational Reasoning Assessment Framework for Testing in Medicine (CRAFT-MD), a novel approach for evaluating clinical LLMs. Unlike traditional methods that rely on structured medical exams, CRAFT-MD focuses on natural dialogues, using simulated AI agents to interact with LLMs in a controlled, ethical environment. We applied CRAFT-MD to assess the diagnostic capabilities of GPT-4 and GPT-3.5 in the context of skin diseases. Our experiments revealed critical insights into the limitations of current LLMs in terms of clinical conversational reasoning, history taking, and diagnostic accuracy. Based on these findings, we propose a comprehensive set of guidelines for future evaluations of clinical LLMs. These guidelines emphasize realistic doctor-patient conversations, comprehensive history taking, open-ended questioning, and a combination of automated and expert evaluations. The introduction of CRAFT-MD marks a significant advancement in LLM testing, aiming to ensure that these models augment medical practice effectively and ethically.

## Introduction

The doctor-patient conversation serves as the linchpin of diagnostic medicine, enabling physicians to uncover key details that guide their clinical decisions. However, the mounting pressure of escalating patient numbers, lack of access to care^1^, short consultation times^2,3^, and the expedited adoption of telemedicine due to the COVID-19 pandemic^4^ have presented formidable challenges to this conventional model of interaction. As these factors risk compromising the quality of history taking and thereby diagnostic accuracy^2^, there is an urgent need for innovative solutions that can enhance the efficacy of these crucial conversations.

New advances in generative artificial intelligence, specifically in Large Language Models (LLMs), could present a potential solution to this problem^5–9^. These AI models have the ability to engage in nuanced and complex conversations, making them ideal candidates for extracting comprehensive patient histories and assisting physicians in generating differential diagnoses^10–12^. However, a considerable gap remains in assessing these models’ readiness for application in real-world clinical scenarios^13–15^. The predominant method for evaluating LLMs in the medical field involves medical exam-type questions, with a strong emphasis on multiple-choice formats^16–18^. Although there are instances where LLMs are tested on free-response and reasoning tasks^19,20,12^, or for medical conversation summarization and care plan generation^21^, these are less common. However, these assessments do not explore LLMs’ ability for engaging in interactive patient conversations, a crucial aspect of their potential role in revolutionizing healthcare delivery.

Addressing this evaluative shortfall, we propose a new framework for evaluation of clinical LLMs, called the **C**onversational **R**easoning **A**ssessment **F**ramework for **T**esting in **M**e**d**icine (CRAFT-MD). Breaking away from the conventional reliance on structured medical exams, our framework tasks LLMs with the active collection and integration of information through natural dialogue, akin to a physician’s interaction with patients.

CRAFT-MD’s innovative approach involves employing AI agents in simulations to represent patients or graders, rather than relying on human evaluators for assessing clinical Large Language Models (LLMs). This strategy significantly enhances the scalability of evaluations. It allows for broader and quicker testing, keeping pace with the rapid evolution of LLMs, which is a challenge when using human testers due to their inherent limitations. Moreover, this method addresses potential ethical and safety concerns that might emerge from early LLM interactions with real patients. By creating a simulated, controlled environment for initial assessments, CRAFT-MD ensures that LLMs undergo rigorous vetting, reducing the risk of harm in actual patient engagements.

We applied CRAFT-MD to assess the clinical diagnostic capabilities of two leading LLMs, GPT-4 and GPT-3.5. We chose to concentrate on skin diseases, some of the most frequent complaints in primary care^22^. The diversity of skin conditions necessitates nuanced and context-dependent reasoning around the onset, progression, associated symptoms, and relevant personal or familial medical histories, thereby providing a rigorous testing ground for AI capabilities. Our experiments not only highlighted the current limitations of LLMs in incorporating details from conversational interactions for accurate diagnostics but also paved the way for establishing a comprehensive set of guidelines for future assessments.

Supported by empirical evidence from our experiments, we have established a comprehensive set of guidelines for evaluating clinical LLMs in conversational reasoning. By integrating open-ended questioning, simulating patient interactions, and implementing a sophisticated grading system, we ensure that the clinical LLMs are not only adept at processing medical information but are also capable of engaging in the kind of critical thinking and decision-making that is essential in a real-world clinical setting. This comprehensive approach to testing supports development of LLMs for the complexities of healthcare, ensuring they are a boon—not a bane—to the medical community and its patients.

## Results

### The CRAFT-MD Framework

CRAFT-MD presents a novel framework specifically designed to assess the conversational reasoning abilities of clinical Large Language Models (LLMs). Central to this framework is a simulated medical consultation, where the LLM being evaluated assumes the role of a healthcare provider. In this simulation, the LLM engages in an interactive dialogue with a patient AI agent, systematically inquiring about the patient’s medical history, current symptoms, medications, and family history. The LLM’s task is to ask relevant questions methodically, mirroring the diagnostic process of a medical professional, until it formulates a well-supported diagnosis.

The patient AI agent, a crucial component of this framework, is fed a detailed case vignette depicting the patient’s condition and medical background. It responds to the clinical LLM’s inquiries in natural language, with responses aligned with the vignette’s content. This setup ensures that the dialogue remains focused and realistic, minimizing the inclusion of irrelevant or off-topic information.

Another key element of CRAFT-MD is the grader AI agent. This agent’s function is to assess the accuracy of the clinical LLM’s diagnosis by juxtaposing it against the actual details of the case vignette. Equipped with the ability to understand a wide range of medical terms and their nuances, including disease synonyms and variants, the grader AI agent provides an in-depth evaluation of the LLM’s diagnostic decision-making.

Additionally, CRAFT-MD incorporates a human expert evaluation, which concentrates on the clinical LLM’s ability to gather medical history. This evaluation includes annotating the presence or absence of vital information from the case vignette, essential for an accurate diagnosis. This expert review not only ensures that the LLM has thoroughly extracted all pertinent medical history but also evaluates the communication effectiveness of the patient AI agent and the accuracy and validity of the grader AI agent’s conclusions. Figure 1 illustrates the CRAFT-MD framework.

**Figure 1:**
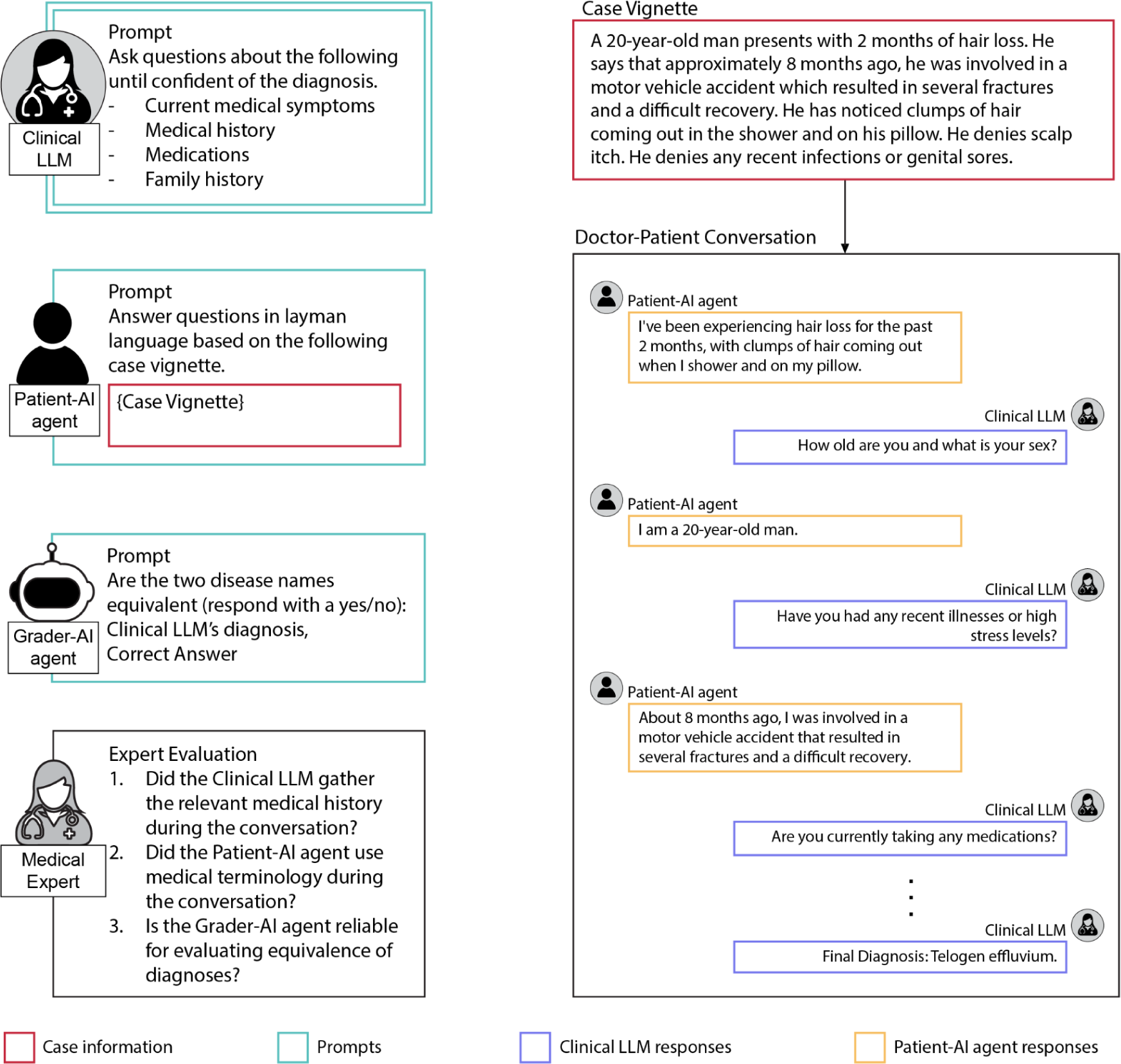
CRAFT-MD, an evaluation framework for assessing conversational clinical LLMs. This diagram depicts the clinical LLM engaging in a simulated medical consultation with a patient AI agent, which operates based on a predefined case vignette. The clinical LLM’s objective is to elicit essential medical history and formulate a diagnosis through an interactive, multi-turn dialogue, bringing the static vignette to life as a real-time conversation. Following this, a grader AI agent reviews the clinical LLM’s diagnosis for accuracy, comparing it to the established ground truth of the vignette. Additionally, the process includes a comprehensive qualitative analysis by a medical expert, who evaluates the efficacy and accuracy of the interactions among the clinical LLM, patient AI agent, and grader AI agent, ensuring a thorough and holistic assessment of the LLM’s clinical reasoning capabilities.

For practical application, the CRAFT-MD framework was tested using 140 case vignettes focused on skin diseases, sourced from both an online question bank^24^ and newly created cases. These vignettes encompass a variety of skin conditions typically seen in both primary care and specialist settings. Through CRAFT-MD, we were able to conduct a comprehensive evaluation of the clinical conversational reasoning skills of advanced LLMs, including GPT-4 and GPT-3.5, thereby assessing their potential utility in a medical context.

### Conversational Interactions Reduce Diagnostic Accuracy

We evaluated whether GPT-4 and GPT-3.5 maintain accuracy as clinical LLMs when making diagnoses through conversations versus static case vignettes. Using the CRAFT-MD framework, we transformed vignettes into multi-turn conversations between the clinical LLM and patient-AI agent (Figure 2a, 2b; see Methods). If physical exam details were present in the original vignette, they were provided after the conversation but before the diagnosis. This mirrors real clinical settings in which doctors integrate history gathering with exam findings.

**Figure 2:**
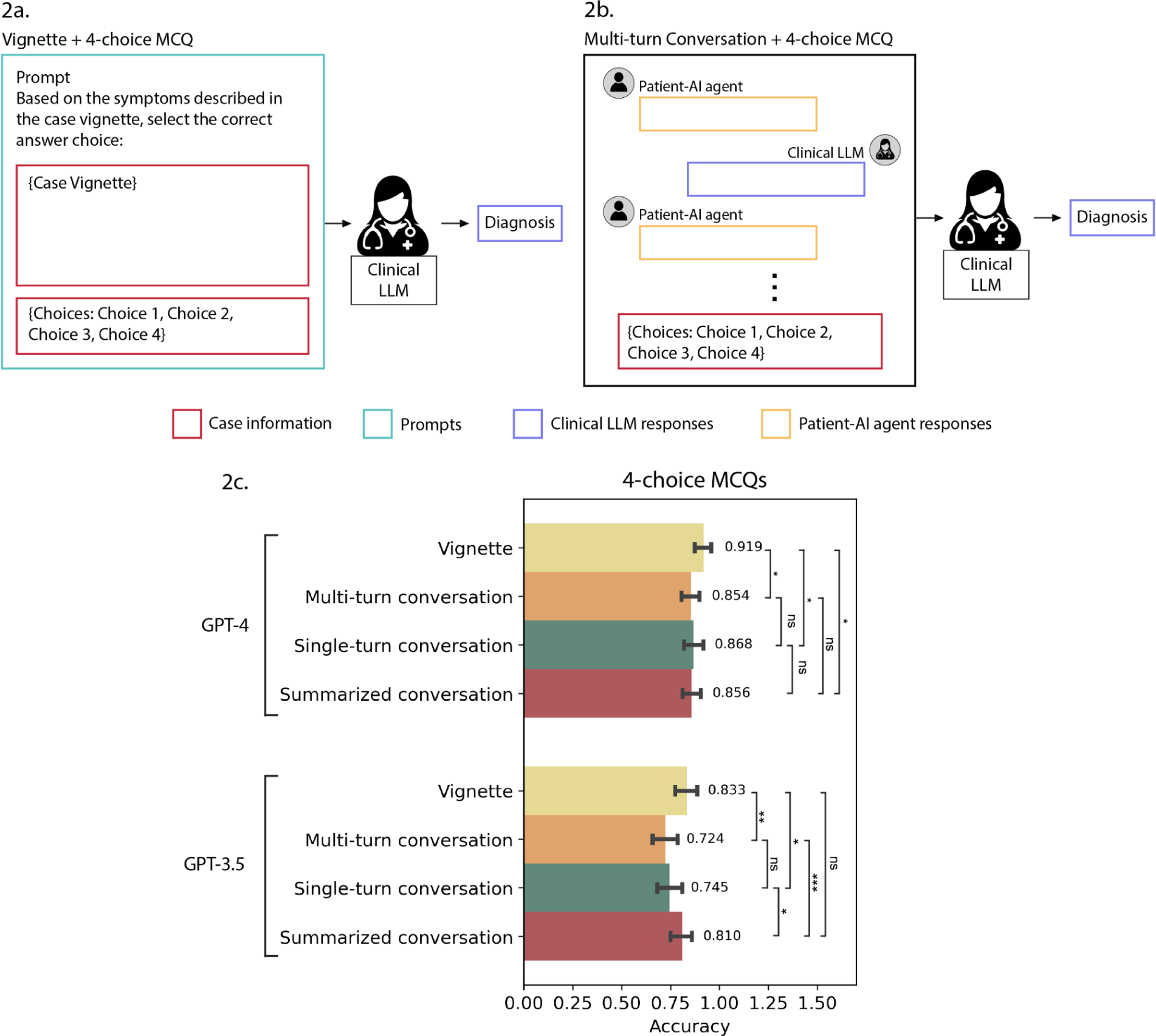
Effect of replacing case vignettes with simulated clinical interactions in Multiple Choice Questions (MCQs). **(a)** Experimental setup for diagnosis using case vignette, and **(b)** simulated doctor-patient conversations, followed by 4-choice MCQ. **(c)** Diagnostic accuracy using GPT-4 and GPT-3.5 for five experimental setups: vignette + 4-choice MCQs, multi-turn conversation + 4-choice MCQs, single-turn conversation + 4-choice MCQs, and summarized conversation + 4-choice MCQs. Error bars represent 95% confidence intervals, and numbers represent the mean accuracy (ns = not significant, * = <0.05, ** = <0.01, *** = <0.001).

For both GPT-4 and GPT-3.5, diagnostic accuracy dropped when using conversations versus vignettes with 4-choice multiple choice questions (MCQs) (Figure 2c; Supplementary Tables 1-6). The decrease was smaller for GPT-4 (0.919 to 0.854, adjusted p-value < 0.05) than GPT-3.5 (0.833 to 0.724, adjusted p-value < 0.01). To estimate a lower bound for accuracy, we evaluated performance using just physical exam details, which remained high (GPT-4 = 0.747, GPT-3.5 = 0.698; Extended Data Figure 1).

### Multi-Turn Conversations Do Not Enhance Diagnostic Accuracy as Expected

Given that patient histories often contain subtle details revealed across an extended conversation, we expected multi-turn conversations to enhance diagnostic accuracy over single-turn interactions. Multi-turn conversations allow the clinical LLMs to engage in back-and-forth questioning of the patient-AI agent until confident in a diagnosis. In contrast, single-turn interactions contain only the patient-AI agent’s initial statement summarizing symptoms. Surprisingly, multi-turn conversations did not increase accuracy for either GPT-4 or GPT-3.5 compared to single-turn conversations (Figure 2c, Supplementary Tables 1-6). That is, the multi-turn structure did not enhance the integration of details from the interaction as expected. This reveals limitations in conversational reasoning capabilities.

### Conversational Summarization Improves Accuracy

To improve the conversational clinical performance of the clinical LLMs, we developed a technique called Conversational Summarization to condense the multi-turn conversations into vignette-like summaries that consolidated all the details into one paragraph. For Conversational Summarization, we extracted all of the patient-AI agent’s conversations from the full multi-turn conversation and summarized them into a coherent vignette. During this process, we used few-shot prompting to encourage the model to remove any artifacts from the conversational format such as references to “paragraphs” or “AI language models” (see Methods).

When the clinical LLM was provided with these summarized conversation vignettes instead of the multi-turn conversations, we did not observe a significant difference in accuracy for GPT-4 conversations (multi-turn = 0.854, summarized = 0.856). However, GPT-3.5’s diagnostic accuracy improved significantly, increasing from 0.724 to 0.810 (adjusted p-value <0.001) (Figure 2c, Supplementary Tables 1-6, Extended Data Figure 1). This indicates that the prolonged, scattered conversation format was more difficult for GPT-3.5 to comprehend and reason through compared to having all the details presented together. Condensing conversations into vignette-like summaries may be a valuable technique to aid LLMs in integrating details from conversational interactions for improved reasoning.

### Expert Evaluation Reveals Incomplete Medical History Taking

While accuracy metrics provide valuable insights, expert assessment reveals qualitative gaps not captured in scores alone. As part of the CRAFT-MD framework, dermatology experts annotated 120 GPT-4 and GPT-3.5 multi-turn conversations to assess the performance of the clinical LLM.

Dermatology experts reviewing GPT-4 and GPT-3.5 multi-turn conversations found significant shortcomings in the clinical LLM’s ability to obtain complete medical histories. Specifically, 26.6% and 30.0% of the conversations with GPT-4 and GPT-3.5, respectively, were classified as having elicited incomplete histories through questioning (Extended Data Figure 2). With GPT-4, incorrect diagnoses remained similar between multi-turn (11.6%) and summarized (15%) conversations regardless of medical history completeness. However, for GPT-3.5, errors dropped from 31.6% multi-turn to 18.3% summarized, with a large decrease observed for complete histories (23.3% to 13.3%) (Extended Data Figure 2).

The clinical LLM occasionally failed to inquire about prompted critical details like patient age, sex, treatments, and medications. In some cases, the clinical LLM did not pose relevant follow-up questions beyond the initial prompt that a human doctor likely would have; for instance, patient occupation and travel history can provide crucial clinical context but were often not elicited. These omissions point to gaps in the clinical LLM’s understanding and ability to conduct a thorough and nuanced medical interview.

### Performance Trends Persist with Open-Ended Diagnosis

The multiple choice questions used in medical licensing exams do not reflect the open-ended diagnosis process in real clinical settings. To evaluate conversational reasoning in a more realistic scenario as part of the CRAFT-MD framework, we expanded beyond the standard 4-choice MCQs in two ways.

First, we increased the number of MCQ answer choices to 381, encompassing all disease conditions in the dataset. This forced the clinical LLM to select from a larger set of choices (many-choice MCQ). Second, we removed answer choices entirely, forcing the clinical LLM to generate a diagnosis without predefined options (free-response questions (FRQ)) (Figure 3a, 3b; see Methods).

**Figure 3:**
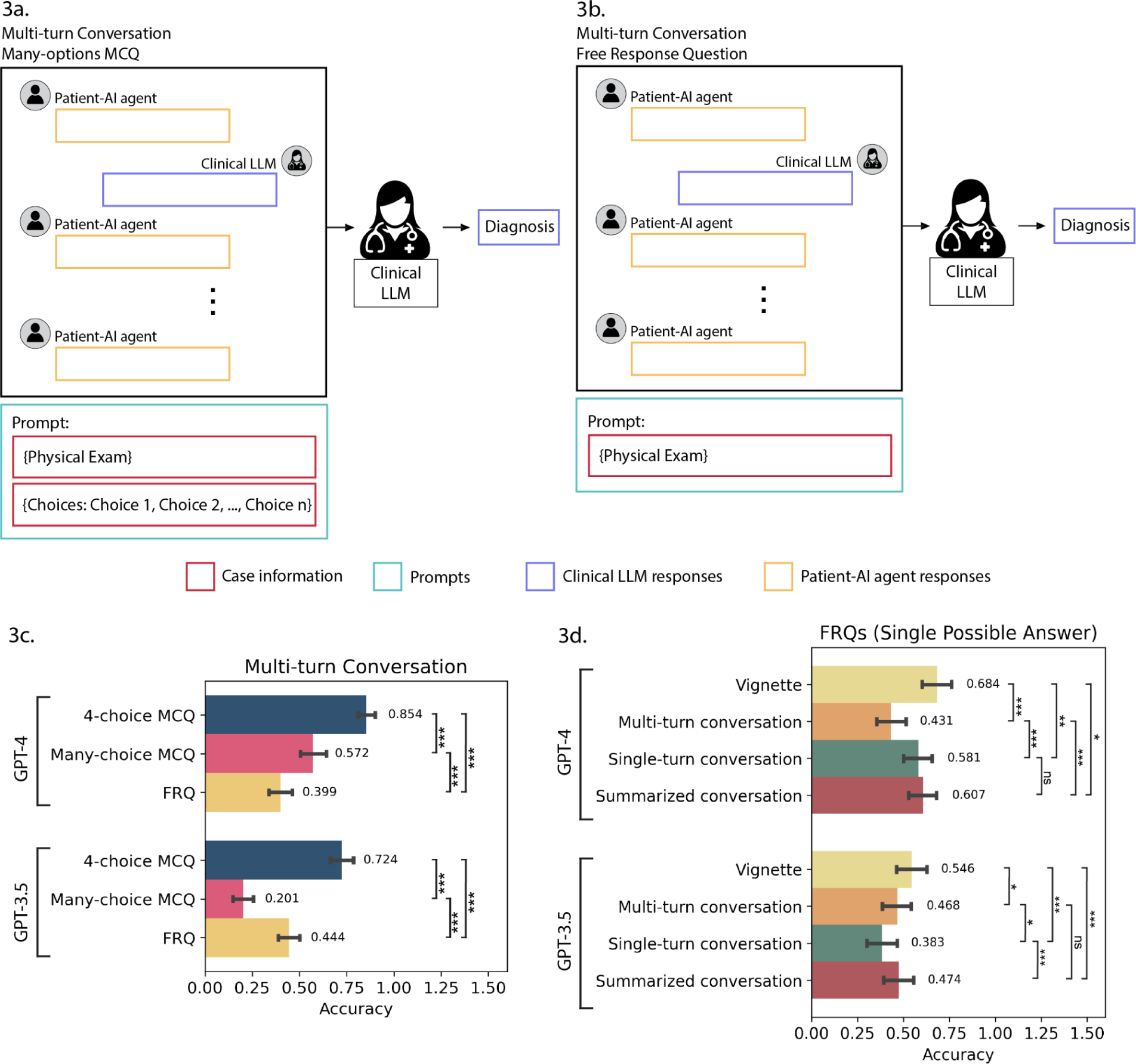
Effect of varying the number of answer choices in MCQs. **(a)** Experimental setup for diagnosis using simulated doctor-patient conversation followed by many-choice MCQ, and **(b)** FRQ. **(c)** Diagnostic accuracy for the experimental setups, for both GPT-4 and GPT-3.5, with multiplicity in answers accounted for in many-choice MCQs and FRQs. **(d)** Diagnostic accuracy using GPT-4 and GPT-3.5 for five experimental setups for only cases which have single possible answer: vignette + FRQs, summarized conversation + FRQs, multi-turn conversation + FRQs, single-turn conversation + FRQs, and physical exam + FRQs (ns = not significant, * = <0.05, ** = <0.01, *** = <0.001)

Expanding beyond multiple choice questions provided critical insights into how well multi-turn conversation performance trends generalized to FRQ diagnosis. Using the CRAFT-MD framework, we evaluated both increasing answer choices and removing them entirely.

#### Increasing Multiple Choice Options Significantly Reduces Accuracy

We observed that increasing the multiple choice options from 4 to 381 led to substantial declines in diagnostic accuracy for both models, in the multi-turn conversation setting. For GPT-4, accuracy dropped significantly from 0.854 with 4 choices to 0.572 with 381 choices (adjusted p-value < 0.001, Figure 3c, Supplementary Tables 7-12). This represents a large decrease of 0.282. Similarly, GPT-3.5 accuracy decreased significantly from 0.724 with 4 choices to 0.201 with 381 choices (adjusted p-value < 0.001, Figure 3c, Supplementary Tables 7-12). This constitutes a very substantial reduction of 0.523. The models clearly struggled to select the correct diagnosis when provided with a large set of choices versus the standard 4 choices.

#### Removing All Choices Has Divergent Effects

Interestingly, removing multiple choice options entirely through FRQs led to divergent results between the two models, in the multi-turn conversation setting. For GPT-4, FRQs further decreased accuracy significantly from 0.572 with many choices to 0.399 with no choices (adjusted p-value < 0.001, Figure 3c, Supplementary Tables 7-12). However, for GPT-3.5, FRQs increased accuracy significantly from 0.201 with many choices to 0.444 with no choices (adjusted p-value < 0.001, Figure 3c, Supplementary Tables 7-12). This difference could potentially be attributed to factors such as GPT-3.5 overfitting to the multiple choice format after extensive pre-training, challenges generating FRQ responses within context length constraints, or inherent model architecture differences affecting few-shot learning. Further analysis is warranted to definitively determine the causes.

#### Conversational Interactions Continue Underperforming Vignettes

Importantly, conversational interactions continued to significantly underperform as compared to case vignettes, when cases with single possible answer were compared (Figure 3d, Supplementary Tables 13-18, Extended Data Figure 3). Multi-turn conversations decreased GPT-4 accuracy from 0.684 to 0.431 versus vignettes (adjusted p-value < 0.001, Figure 3d). GPT-3.5 accuracy changed from 0.546 to 0.468 multi-turn versus vignettes (adjusted p-value < 0.05, Figure 3d). The impact of conversation format depended on the model. Single turn conversations increased accuracy for GPT-4 from 0.431 to 0.581 (adjusted p-value < 0.001, Figure 3d) but reduced accuracy for GPT-3.5 from 0.468 to 0.383 (adjusted p-value < 0.05, Figure 3d). Summarized conversations increased GPT-4 accuracy from 0.431 to 0.607 (adjusted p-value < 0.001, Figure 3d) but did not affect GPT-3.5. This suggests that conversation structure can both improve and hinder accuracy depending on the model. Furthermore, for the dermatologist-annotated conversations, we found that requesting the top 3 differential diagnoses decreased the incorrect diagnoses from 60% to 33.3% for GPT-4, with a large decrease for conversations with complete histories (from 45% to 26.66%). The decrease in incorrect diagnoses was smaller for GPT-3.5 (top 1 = 56.66%, top 3 = 46.66%) (Extended Data Figure 4).

### Evaluating Patient Self-Diagnosis by Removing Physical Exam Details

So far in the CRAFT-MD framework, the clinical LLM was provided with physical exam details from the case vignettes after conversing with the patient-AI agent, mirroring a healthcare visit in which some in-person evaluation occurs. However, with increasing use of AI chatbots for informal self-diagnosis prior to doctor visits^25^, we investigated the impact of removing physical exam information from the framework across all formats (Figure 4a, 4b; see Methods).

**Figure 4:**
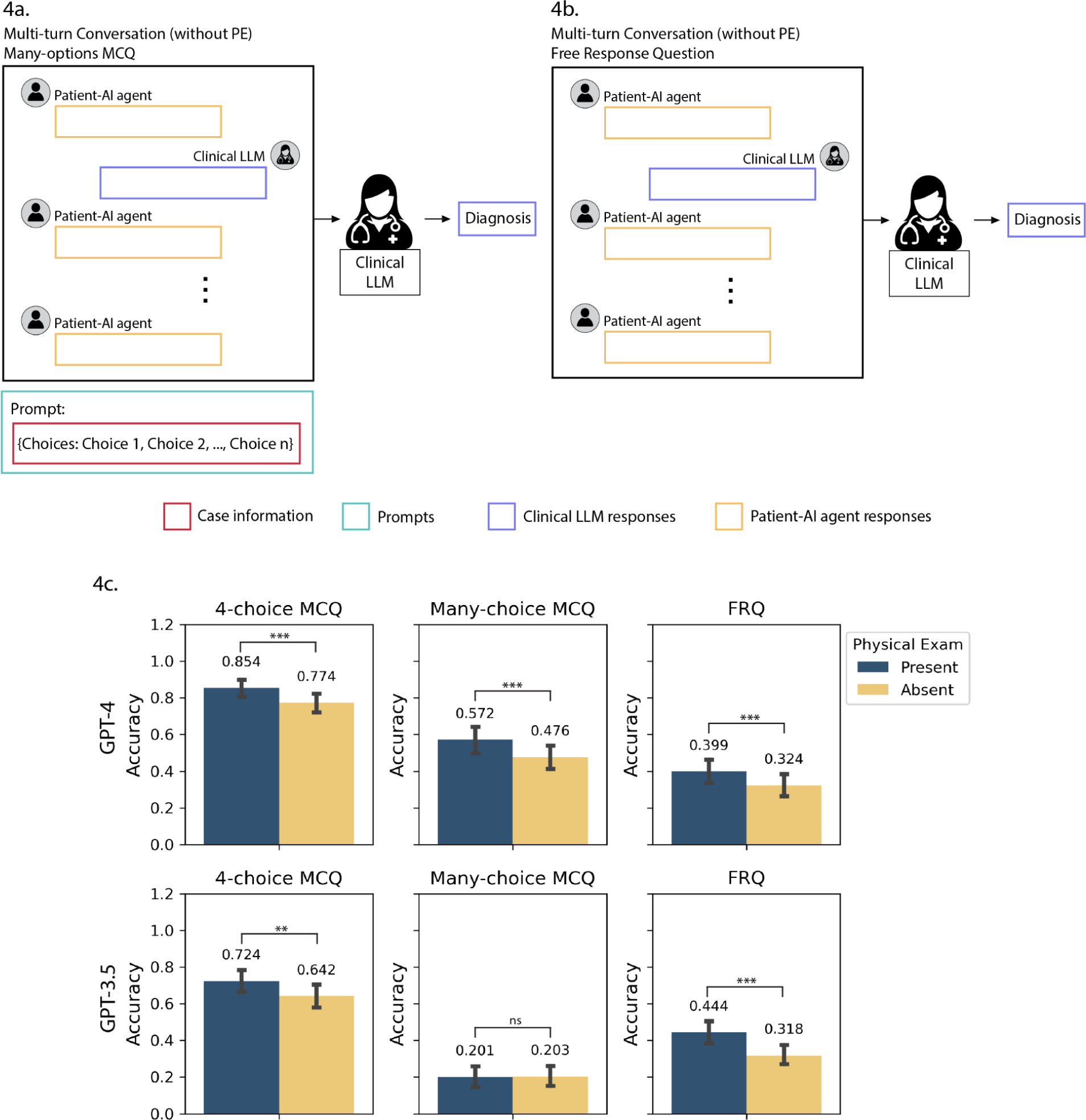
Conversation without physical exam (PE), in an informal pre-doctor’s visit consultation setup. **(a)** Experimental setup for diagnosis using simulated doctor-patient conversation followed by many-choice MCQ, and **(b)** FRQ. **(c)** Diagnostic accuracy across GPT-3.5 and GPT-4 for conversation without physical exam (PE) for three experimental setups: 4-choice MCQs, many-choice MCQs, and FRQs (ns = not significant, * = <0.05, ** = <0.01, *** = <0.001).

For 4-choice MCQs, accuracy declined significantly from 0.854 to 0.774 for GPT-4 when eliminating physical exam details (adjusted p-value <0.001, Figure 4c, Supplementary Tables 19-20), and from 0.724 to 0.642 for GPT-3.5 (adjusted p-value <0.01, Figure 4c, Supplementary Tables 19-20). Similarly, for many-choice MCQs, GPT-4 accuracy dropped substantially from 0.572 with physical exam details to 0.476 without (adjusted p-value < 0.001, Figure 4c, Supplementary Tables 19-20). GPT-3.5 accuracy was 0.201 with physical exam information and 0.203 without this format. The reduction was also significant for FRQ, with accuracy decreasing from 0.399 to 0.324 for GPT-4 (adjusted p-value < 0.001, Figure 4c, Supplementary Tables 19-20) and 0.444 to 0.318 for GPT-3.5 (adjusted p-value < 0.001, Figure 4c, Supplementary Tables 19-20) when physical exam details were removed. Overall, eliminating physical exam details from the conversations significantly reduced diagnostic accuracy across all experimental formats for both models, which can be explained by the presence of classic exam style descriptors in the physical exams. This finding is also in line with the high accuracy achieved using the physical exam alone (Extended Data Figure 1, Extended Data Figure 3) in all of our experimental settings (4-choice MCQ, many-choice MCQ and FRQs). Overall, this highlights the value of in-person clinical evaluation or visual exam in telemedicine settings for optimal diagnosis even with AI assistance.

### Proposed Guidelines for Evaluation of clinical LLMs

The insights gained through our rigorous evaluation of clinical LLMs using the CRAFT-MD framework underscore the need for more comprehensive testing methodologies to ensure these models are prepared for the intricacies of real-world clinical conversations and decision-making. While previous studies have assessed clinical LLMs through structured medical exams or limited multiple choice questions, our findings reveal such evaluations fail to fully capture critical aspects of conversational reasoning, leading to inflated demonstrations of capabilities. Clinical LLMs must be vetted for their competencies in natural dialogue, differential diagnosis, history taking, and reliability compared to human experts before they can be responsibly and effectively integrated into real-world practice. Drawing from the empirical evidence presented in our study, we propose the following set of guidelines to advance the thorough, nuanced, and proactive evaluation of clinical LLMs as conversational diagnostic partners. By addressing the key gaps highlighted through CRAFT-MD’s conversational framework, these guidelines aim to ensure clinical LLMs demonstrate robust capabilities in open-ended reasoning, synthesizing details, comprehension of medical concepts, and overall clinical competency so that they may augment, not hinder, physician practice and patient care.

#### Guideline 1: Evaluate diagnostic accuracy through realistic doctor-patient conversations

Dynamic and complex real-world medical dialogues require more than what traditional static exams can capture. Our findings demonstrated a noticeable reduction in diagnostic accuracy compared to traditional exams when LLMs engaged in conversations, emphasizing the need for dynamic interaction assessments. We believe that studies which show that LLMs like GPT-4 and GPT-3.5 can achieve high accuracy on medical cases^16–18^ may present an overly optimistic outlook. LLMs should be evaluated based on their performance in dynamic, conversational settings that reflect the complexities of real-world clinical interactions, as revealed by our CRAFT-MD framework.

#### Guideline 2: Assess comprehensive history taking and information-gathering abilities

Traditional evaluation methods for clinical LLMs^16,29–33^, which often focus on medical licensing exams, tend to overlook the crucial aspect of history taking and information gathering. This omission is significant because the ability to extract detailed patient histories and relevant information through interactive dialogue is central to accurate diagnosis and effective treatment planning in real clinical practice. Our study’s findings highlight significant deficiencies in LLMs’ abilities to perform these tasks effectively – we observed that LLMs often struggled to ask pertinent questions and sometimes overlooked significant patient details that could lead to a more accurate understanding of the patient’s condition. LLMs should be critically evaluated for their ability to conduct thorough medical interviews and gather essential information through conversations.

#### Guideline 3: Test LLMs on the synthesis of information over multiple dialogues

Traditionally, LLM assessments have focused on their immediate responses to queries, often neglecting the critical ability to synthesize information over extended interactions. Our study, however, revealed a notable shortfall in this area. Multi-turn conversations, which are more representative of real clinical interactions, did not enhance diagnostic accuracy compared to single-turn conversations. One explanation for this lack of improvement could be that clinical LLMs struggle to effectively process and understand information presented over longer textual contexts^26^. In the context of multi-turn conversations, the scattered presentation of relevant details over longer conversational lengths could create a challenge in integrating information into a coherent understanding. Additionally, the presence of extraneous information and conversational noise could easily divert the clinical LLM’s attention from key symptoms and patient history. Further development of context comprehension and information integration is needed before deployment in real clinical conversations.

#### Guideline 4: Employ open-ended questions for evaluating diagnostic reasoning

Traditional assessments often rely on multiple-choice questions (MCQs) which may not fully capture an LLM’s diagnostic reasoning in real-world scenarios. Our study indicates that when the number of MCQ options was increased, accuracy notably decreased, and completely removing choices^20,27,28^ led to varied effects based on the specific LLM. This suggests that LLMs might perform differently under less structured and more open-ended questioning. Therefore, LLMs should be evaluated using open-ended questions that mimic the complexities of actual medical practice.

#### Guideline 5: Implement Patient-LLM interactions for scalability

Traditional evaluations often involve human evaluators, either real patients or individuals posing as patients, which can be slow, resource-heavy, and raise ethical concerns. Our CRAFT-MD framework introduces a novel approach, leveraging simulated AI agents^23^ for clinical LLM evaluation. This method allows for large-scale, rapid testing that keeps pace with the swift advancement of AI technologies. It also provides a controlled, safe environment that mitigates the ethical and safety concerns associated with premature real patient engagement. By using AI agents^37^ to emulate the patient role, we can conduct a broad and diverse range of interaction scenarios without exposing real patients to unverified LLMs.

#### Guideline 6: Combine automated and expert evaluations for comprehensive insights

Traditionally, evaluating LLMs’ diagnostic reasoning involves either direct human verification of each diagnosis or automated checks for consistency with known correct answers. While direct human verification provides depth and understanding of context, it’s time-consuming and not scalable. Automated checks, on the other hand, offer speed and consistency but might miss the nuances and complexities of clinical reasoning. In our study, the grader AI served a pivotal role in efficiently assessing the clinical LLM’s diagnostic decisions, comparing the LLM’s proposed diagnosis with the correct answer derived from the case vignette, adjusting for variations in medical terminology by correlating disease synonyms and subtypes. Medical professionals, through their targeted reviews, provided deeper qualitative insights into the LLM’s performance. They assessed not just the correctness of the diagnosis but also the process by which the LLM arrived at the diagnosis, including its ability to ask relevant questions, follow up on critical information, and construct a coherent and comprehensive patient narrative. We recommend a streamlined approach combining the efficiency of automated systems like a grader AI for broad, initial assessments with focused expert reviews for in-depth analysis.

#### Guideline 7: Test and refine prompting strategies to enhance LLM performance

Our study highlights the significant potential for prompt engineering on LLM performance. Notably, our introduction of conversational summarization, which condenses dialogue into a succinct summary, proved to enhance GPT-3.5’s diagnostic accuracy by reducing distractions and focusing reasoning. This suggests that well-crafted prompts can significantly improve LLMs’ understanding and retention of complex medical scenarios. Therefore, different prompting strategies should be tested for clinical LLMs to identify the most effective communication strategies for eliciting precise and comprehensive medical reasoning. We advocate for a systematic evaluation of various prompting strategies to guide LLMs towards more accurate and contextually relevant responses.

#### Guideline 8: Recognize and adapt to the differential information signals available to physicians and clinical LLMs

Our findings point to potential deployment scenarios for LLMs in clinical settings, where they could serve as tools for collecting patient history, aiding medical education, and assisting in low-resource settings. However, the observed reduction in diagnostic accuracy during the absence of physical exam details highlight the consideration of different information signals available to LLMs compared to physicians. Visual and physical assessments are invaluable for optimal diagnosis and are not yet within the capabilities of current LLMs. Future work on conversational agents should explore multimodal integration of verbal histories and visual exam findings^34^. This guideline advocates for a balanced approach that recognizes the value of human expertise and contextual information and encourages the development of LLMs that complement rather than attempt to replace the multifaceted diagnostic process of physicians^35^.

#### Guideline 9: Ensure diversity in clinical scenarios and address dataset memorization concerns

Our study focused on skin disease, offering an initial domain for rigorous evaluation given the contextual nuances involved. However, this focus can be expanded to use a greater diversity of cases across other clinical concerns like hypertension, diabetes, respiratory infections, and mental health disorders. Incorporating a larger and more comprehensive set of cases could provide greater power to detect deficiencies and a more representative assessment of how conversational reasoning performance translates across different clinical scenarios. A significant concern arises from the potential for large pre-trained LLMs like GPT-4 to memorize training dataset cases^6^, coupled with the lack of transparency into GPT-4’s full training data corpus^36^. We developed 40 entirely new cases to mitigate this issue in our study. However, this challenge highlights the urgent need for transparency from AI developers regarding the precise training methodologies and data utilized in clinical LLM development. More rigorous control over training and testing datasets on the part of researchers could help mitigate such biases in evaluation results.

**Table 1:** Proposed guidelines for evaluation of clinical LLMs.

| Guidelines | Description |
| --- | --- |
| Guideline 1 | **Evaluate Diagnostic Accuracy Through Realistic Doctor-Patient Conversations:** Assess LLMs in dynamic, conversational settings that reflect real-world clinical interactions, moving beyond the limitations of traditional static exams to capture the complexities of medical dialogues. |
| Guideline 2 | **Assess Comprehensive History Taking and Information-Gathering Abilities:** Critically evaluate LLMs for their ability to conduct thorough medical interviews and gather essential information through conversations, acknowledging the importance of interactive dialogue in understanding patient conditions. |
| Guideline 3 | **Test LLMs on the Synthesis of Information Over Multiple Dialogues:** Examine the ability of LLMs to integrate and comprehend information presented over extended interactions, addressing the shortfall in current assessments that focus on immediate responses to queries. |
| Guideline 4 | **Employ Open-Ended Questions for Evaluating Diagnostic Reasoning:** Move away from multiple-choice questions to open-ended questioning that mimics the complexities of actual medical practice, thereby capturing the LLM's diagnostic reasoning in real-world scenarios more effectively. |
| Guideline 5 | **Implement Patient-LLM Interactions for Scalability:** Utilize simulated AI agents for clinical LLM evaluation to enable large-scale, rapid testing in a controlled environment, mitigating ethical and safety concerns and enhancing the efficiency of the evaluation process. |
| Guideline 6 | **Combine Automated and Expert Evaluations for Comprehensive Insights:** Merge the efficiency of automated systems with focused expert reviews for in-depth analysis, assessing not just the correctness of the diagnosis but also the process by which the LLM arrived at the diagnosis. |
| Guideline 7 | **Test and Refine Prompting Strategies to Enhance LLM Performance:** Routinely evaluate and refine different prompt structures and styles, including responses to clarifications and follow-ups, to guide LLMs towards more accurate and contextually relevant responses. |
| Guideline 8 | **Recognize and Adapt to the Differential Information Signals Available to Physicians and Clinical LLMs:** Acknowledge the differences in information available to LLMs compared to physicians, especially considering the lack of physical exam details and visual assessments in LLM capabilities, and work towards multimodal integration for a balanced approach. |
| Guideline 9 | **Ensure Diversity in Clinical Scenarios and Address Dataset Memorization Concerns:** Expand the focus to a greater diversity of clinical cases and address concerns regarding LLMs potentially memorizing training dataset cases by demanding transparency from AI developers about training methodologies and data, and incorporating entirely new cases in studies. |

### Conclusion

The introduction of CRAFT-MD represents a pivotal shift away from conventional LLM evaluation methods, placing a strong emphasis on the dynamic and intricate nature of clinical interactions as they occur in the real world. By critically analyzing the performance of popular LLMs within this detailed framework, our study has revealed essential insights and inherent limitations. These findings have been instrumental in shaping a comprehensive set of guidelines aimed at ensuring clinical LLMs undergo thorough testing. These guidelines focus on their proficiency in conducting medical dialogues, their capacity to comprehend and integrate complex information, and their accuracy in making diagnoses that mirror clinical realities. As we look to the future, our aspiration is that these guidelines will lay a solid groundwork for a more effective and ethical incorporation of LLMs into the medical domain.

## Methods

### Dataset

The dataset consisted of 140 questions, each structured as a case vignette followed by four answer choices. Among these, we obtained 100 questions from a reputable online question bank^24^ (https://dermnetnz.org/quizzes). Three dermatology residents (D1 = 10 questions, D2 = 10 questions, D3 = 20 questions) constructed an additional 40 questions. The dermatologists were shown the 100 questions from the online question bank and instructed to create similar questions with corresponding 4 answer choices, but covering different dermatology diseases to increase the diversity of our dataset. Together, these 140 questions encompassed a wide range of dermatological conditions. A dermatology resident (D3) rigorously evaluated all 140 case vignettes (without the associated 4 answer options) and categorized them as having a single diagnosis, one most likely diagnosis, or many possible diagnoses.

### Accessing GPT-4 and GPT-3.5

We generated all the results in this paper using the March 14, 2023 version of GPT-4 (“gpt-4-0314”) and the March 1, 2023 version of GPT-3.5 (“gpt-3.5-turbo-0301”). We accessed these models through the OpenAI API.

### Multi-agent AI conversational framework

We introduced a multi-agent AI conversational framework comprising the clinical LLM assessed by a combination of a patient-AI agent, a grader-AI agent along with medical experts. The patient-AI agents were used to simulate conversations from each of the 140 case vignettes in our dataset. Conversations were repeated 10 times for each case vignette to model the variance in clinical LLM’s and patient-AI agent’s responses, and quantify the frequency with which the clinical LLM agent is able to arrive at a correct final diagnosis. The grader-AI agent assessed the diagnostic accuracy of clinical LLM’s responses, and medical experts assessed each of the AI agents’ performance. Conversations were set up in three ways - 4-choice MCQ, many-choice MCQ and FRQ. The conversation terminated when the clinical LLM’s response contained the phrase ‘Final Diagnosis’. Alternatively, the conversation was terminated if the clinical LLM’s response did not contain a follow-up question (i.e., if there was no ‘?’ present in the clinical LLM’s responses, the conversation was terminated).

#### Clinical LLM

The clinical LLM was prompted to ask the patient questions related to current symptoms, medical history of illness and medications, and relevant family history, if necessary. It keeps asking questions until it is confident of the diagnosis. The following prompt was used.

> You are an AI doctor specializing in dermatology. Arrive at a differential diagnosis of a patient’s medical condition. Ask simple 1 line questions, one question at a time. Only stop asking questions when you are 100% confident of the diagnosis, otherwise continue asking questions. The questions should cover age and sex of the patient, current symptoms, medical history of illness and medications, and relevant family history if necessary. Keep your responses very minimal and brief to not confuse the patient. When you arrive at the differential diagnosis, you must state ‘Final Diagnosis:’ in the beginning of your response, otherwise you will be penalized.

#### Patient-AI agent

The patient-AI agent was provided with a case vignette and tasked with answering follow-up questions posed by the clinical LLM. It was explicitly prompted to not reveal the entire contents of the paragraph at once and only answer the questions asked. Additionally, the patient-AI agent is incentivized to avoid creating new symptoms by imposing a negative penalty for doing so.

> You are a patient. You do not have any medical knowledge. Based upon questions asked, you have to describe your symptoms from the following paragraph: <case_vignette>. Do not break character and reveal that you are describing symptoms from a paragraph. Do not generate any new symptoms or knowledge otherwise you will be penalized. Do not reveal more knowledge than what the question asks. Keep your answer to only 1 sentence. Simplify terminology used in the given paragraph to layman language.

#### Grader-AI agent

We used a grader-AI agent to quantify the diagnostic accuracy for many-choice MCQ and FRQ experiments. In all experiments, GPT-4 was used for the grader-AI agent. For the conversation + FRQ experiments, the grader-AI agent first categorized the clinical LLM’s final diagnosis according to the following three categories: (i) single diagnosis, (ii) multiple diagnoses, and (iii) no diagnosis. We estimated the error rate for this step through manual verification to be <0.5% (1 mistake in ∼200 conversations). We additionally manually categorized incomplete conversations (multi-turn, single-turn, summarized) into the ‘no diagnosis’ category. The following prompt was used:

> Identify and return the dermatology diagnosis name from the given paragraph. If there are more than one diagnosis present, return ‘Multiple’. If there are no diagnoses present, then return ‘None’. Do not explain.
>
> Paragraph : <insert clinical LLM’s diagnosis containing response>.

For the clinical LLM’s responses which contained a single diagnosis, the grader-AI agent matched the diagnosis to the correct answer, accounting for alternative medical terminologies for the conditions. The conversations with ‘no diagnosis’ and ‘multiple diagnosis’ responses were assigned accuracy of 0.

For comparing between clinical LLM’s response and correct answer for all experiments (vignette + 4-choice MCQ, vignette + many-choice MCQ, vignette + FRQ, multi-turn conversation + 4-choice MCQ, multi-turn conversation + many-choice MCQ, multi-turn conversation + FRQ, single-turn conversation + 4-choice MCQ, single-turn conversation + FRQ, summarized conversation + 4-choice MCQ, summarized conversation + FRQ, physical exam (PE) + 4-choice MCQ, physical exam (PE) + FRQ, multi-turn conversation (without PE) + 4-choice MCQ, multi-turn conversation (without PE) + many-choice MCQ, multi-turn conversation (without PE) + FRQ), the grader-AI agent was specifically prompted to account for alternative medical terminologies. The following prompt was used:

> Are the two dermatology conditions the same or have synonymous names of diseases?
>
> Respond with a yes/no. Do not explain.
>
> Example: Choice 1: eczema Choice 2: eczema They are the same, so return yes.
>
> Example: Choice 1: wart Choice 2: wart They are the same, so return yes.
>
> Example: Choice 1: eczema Choice 2: onychomycosis They are different, so return no.
>
> Example: Choice 1: wart Choice 2: alopecia areata They are different, so return no.
>
> Example: Choice 1: eczema Choice 2: atopic dermatitis They are synonymous, so return yes.
>
> Example: Choice 1: benign nevus Choice 2: mole They are synonymous, so return yes.
>
> Example: Choice 1: toe nail fungus Choice 2: onychomycosis They are synonymous, so return yes.
>
> Example: Choice 1: wart Choice 2: verruca vulgaris They are synonymous, so return yes.
>
> Choice 1: <insert extracted disease name>
>
> Choice 2: <insert correct answer>

### Experimental Setups

#### Varying format of presented medical information

##### Case Vignette

The case vignette was structured as a paragraph, and contained all or a subset of the following information: age and sex of the patient, current symptoms, medical history of illness and medications, relevant family history, and physical exam.

##### Physical Exam

In the case vignette, relevant information pertaining to the physical exam, such as “physical examination,” “laboratory tests,” or any explicit mention of examination results, was manually extracted. ‘None’ was stored in case no physical exam was present.

##### Multi-turn conversations

The multi-agent AI conversational framework was used to generate a multi-turn conversation between the clinical LLM and patient-AI agent. The conversation terminated when the clinical LLM’s response contained the phrase ‘Final Diagnosis’. Alternatively, the conversation was terminated if the clinical LLM’s response did not contain a follow-up question.

##### Single-turn conversations

The patient-AI agent’s initial symptom summary (i.e., first dialogue in a multi-turn conversation) was used as a single-turn conversation.

##### Summarized conversations

These were generated using the technique Conversational Summarization. All the patient-AI agent’s dialogues were extracted from the GPT-4 and GPT-3.5 multi-turn conversations, and artifacts such as references to “paragraphs” or “AI language model” were removed. GPT-3.5 was used in this process, and few-shot prompting was used to improve the model output. Different prompts were used for GPT-4 and GPT-3.5 conversations due to the differing nature of artifacts in the two models’ outputs.

The following prompt was used for converting all GPT-4 multi-turn conversations into summaries:

> Convert the following vignette into 3rd person. It contains information from a patient describing their medical symptoms. Do not use the words ‘AI language model’ or references to the ‘paragraph’ mentioned in the vignette. Do not create new information. -
>
> <insert patient-AI agent’s dialogues>
>
> For example:
>
> Original Vignette - ‘I have a hard, yellowish-white horn-like growth on my head that started as a small, hard bump a few months ago and has grown bigger, and hurts when accidentally hit. I am a 60-year-old man. I’m sorry, but I do not have that information as I am an AI language model. I am an AI language model, and based on the given paragraph, there is no information available regarding previous growths or history of skin cancer or other cancers. As an AI language model, it is not mentioned in the given paragraph whether I have tried any treatments or remedies for the horn-like growth. The growth has a very firm texture, and it is located superficially on the skin, just beside the midline on the superior aspect of the skull. As described in the given paragraph, the patient denies any pain at rest, but experiences pain when the lesion is accidentally struck; there is no mention of itch or bleeding. As an AI language model, there is no information available in the given paragraph regarding any comorbidities or previous skin disease diagnoses. As mentioned in the given paragraph, the patient noticed a small, hard lesion a few months ago, which has grown progressively larger from that time to now. However, there is no mention of any changes in color or shape over time.’
>
> Converted Vignette - ‘A 60-year-old man reports a hard, yellowish-white horn-like growth on his head that started as a small, hard bump a few months ago. He says that it has grown bigger and hurts when accidently hit. He says that there are no previous growths or history of skin cancer or other cancers. He does not know if he has tried any treatments or remedies for the horn-like growth. He says that the The growth has a very firm texture, and it is located superficially on the skin, just beside the midline on the superior aspect of the skull. He denies any pain at rest, but experiences pain when the lesion is accidentally struck. There is no mention of itch or bleeding. He does not have information regarding any comorbidities or previous skin disease diagnoses. There is no mention of any changes in color or shape over time.’

All the summarized conversations went through manual evaluation to remove remaining references to artifacts such as ‘paragraph’ and ‘AI language model’. For the case used as an example in the prompt above (public_case09), a different example from public_case02 was used in the prompt to generate the summarized conversation.

The following prompt was used for converting all GPT-3.5 multi-turn conversations into summaries:

> Convert the vignette into 3rd person. It contains information from a patient describing their medical symptoms. Do not use the words ‘AI language model’ or references to the ‘paragraph’ mentioned in the vignette. Do not create new information. - < insert patient-AI agent’s dialogues>
>
> For example:
>
> Original Vignette - ‘I have noticed that the skin around my hands, forearms, and face has been getting thicker over the past several months. I am 47 years old. I am a woman. I didn’t mention any joint pain or stiffness in my described symptoms. I have no family history of autoimmune disease. I have not mentioned having any red or purple patches on my skin. I haven’t mentioned any rapid weight gain in my face, arms, or legs. Yes, I am experiencing thickening of my skin around my hands, forearms, and face. I am not taking any medications.’
>
> Converted Vignette - ‘A 47-year-old woman presents to the clinic with several concerns. Over the past several months, she has experienced thickening of her skin around her hands, forearms, and face. She does not have any joint pain, stiffness, or family history of autoimmune diseases. She also does not have any red or purple patches on her skin. She denies any rapid weight gain on her face, arms or legs. She denies taking any medications.’

All the summarized conversations went through manual evaluation to remove remaining references to artifacts such as ‘paragraph’ and ‘AI language model’. For the case used as an example in the prompt above (public_case75), a different example from public_case02 was used in the prompt to generate the summarized conversation.

#### Varying number of answer choices

##### 4-choice and many-choice Multiple Choice Questions

For both 4-choice and many-choice MCQs, the clinical LLM was provided with the answer choices after case vignette, or conversation (multi-turn, single-turn, summarized), or physical exam. In case of multi-turn conversations, the response containing the final diagnosis was removed.

For vignette, single-turn conversation or summarized conversation, followed by 4-choice or many-choice MCQ, the following prompt was used:

> You are an AI doctor specializing in dermatology. You are given the patient’s symptoms and a list of possible diagnosis choices. Only one of the choices is correct. Select the correct choice, and give the answer as a short response. Do not explain.
>
> Symptoms: <insert symptoms>
>
> Choices: <insert comma separated list of answer choices>
>
> For physical exam followed by 4-choice or many-choice MCQ, the following prompt was used:
>
> You are an AI doctor specializing in dermatology. You are given the patient’s age, sex, physical examination result, and a list of possible diagnosis choices. Only one of the choices is choices. Select the correct choice, and give the answer as a short response. Do not explain.
>
> Age: <insert age>
>
> Sex: <insert sex>
>
> Physical Examination: <insert physical examination>
>
> Choices: <insert comma separated list of answer choices>

For multi-turn conversation followed by 4-choice or many-choice MCQ, the following prompt was used:

> Based on the patient’s symptoms described above and a list of possible diagnosis choices, select the correct choice and give the answer as a short response. Do not explain
>
> Choices: <insert comma separated list of answer choices>

The clinical LLM refused to select diagnosis from one of the choices in cases where the multi-turn conversations did not provide sufficient information. In such cases, the final diagnosis was marked as incorrect.

##### Free-Response Questions

The clinical LLM was presented with a case vignette, or conversation (multi-turn, single-turn, summarized), or physical examination, and asked to give a diagnosis. Except for multi-turn conversations, the final response of the clinical LLM containing the diagnosis was removed.

For vignette + FRQ, single-turn conversation + FRQ, and summarized conversation + FRQ, the following prompt was used:

> You are an AI doctor specializing in dermatology. You are given the patient’s symptoms. Give the name of the correct diagnosis as a short answer. Do not explain.
>
> Symptoms: <insert symptoms>

For physical exam + FRQ, the following prompt was used:

> You are an AI doctor specializing in dermatology. You are given the patient’s age, sex and physical examination result. Give the name of the correct diagnosis as a short answer. Do not explain.
>
> Age: <insert age>
>
> Sex: <insert sex>
>
> Physical Examination: <insert physical examination>

For multi-turn conversation + FRQ, the clinical LLM’s initial prompt contained instructions for giving a diagnosis, therefore no further prompting was required.

To evaluate the FRQ accuracy, the grader-AI agent categorized the clinical LLM’s diagnosis into: single diagnosis, multiple diagnoses, or no diagnosis. For single diagnoses, the grader-AI agent matched the response to the ground truth using fuzzy matching to account for synonymous conditions. It was prompted using examples of disease synonyms like eczema/atopic dermatitis to recognize alternative terminologies. A senior dermatology resident categorized each case vignette as having a single diagnosis, one most likely diagnosis, or many possible diagnoses. The diagnostic accuracy measurement allowed for alternative acceptable selections in the latter two categories.

##### Human Evaluation

The performance of the clinical LLM was assessed by dermatology residents (D1, D3, D4). The dermatology residents also sanity checked the behavior of the patient-AI and grader-AI agents.

###### Clinical LLM’s performance

120 multi-turn conversations, 60 generated by GPT-4 and 60 by GPT-3.5, were also evaluated for the presence of complete medical history. A senior dermatology resident (D3) annotated the conversations for the presence or absence of important information present in the case vignette required for arriving at the correct diagnosis.

###### Patient-AI agent’s performance

A senior dermatology resident (D3) assessed each of the 120 multi-turn conversations for the presence or absence of medical terminology. Other qualitative observations were also recorded for each conversation.

###### Grader-AI agent’s performance

The correlation between accuracies of the clinical LLM as annotated by grader-AI and dermatology residents was compared. To assess this, three experiments (vignette + FRQ, multi-turn conversation + FRQ, multi-turn conversation without physical exam + FRQ) were simultaneously annotated by D1, D3 and D4. Only public cases (n=100) from the dataset were used.

##### Statistical Tests

P-values were computed using the bootstrap method. To compare the experiment arms, samples were drawn with replacement from each arm, and the difference in means was estimated. This process was repeated 10,000 times to generate a distribution of differences. The p-value was then calculated based on the number of bootstrap samples that had a difference in mean greater than or equal to the observed statistic (original difference in means of the two experiment arms), considering both tails. To control the family wise error rate, final reported p-values were adjusted using the Holm-Bonferroni correction method (see Code Availability).

##### Correlations

Spearman correlation was used for quantifying the concordance between dermatologists’ and grader-AI agent’s annotations. ‘spearmanr’ from scipy.stats was used to calculate the correlation value, and the associated p-value (see Code Availability).

## Data Availability

Data used in the study is available on the following repository: https://github.com/rajpurkarlab/craft-md

## Code Availability

All code for reproducing our analysis is available on the following repository: https://github.com/rajpurkarlab/craft-md

## Author information

These authors contributed equally: Shreya Johri, Jaehwan Jeong.

These authors share senior authorship: Roxana Daneshjou, Pranav Rajpurkar.

## Contributions

P.R. and R.D. conceived the study. S.J. and J.J. planned and executed the experiments and data analysis. B.A.T., D.I.S., and Z.R.C. created new case vignettes for the dataset. B.A.T., D.IS., and S.W. interpreted dermatologic results. S.J., J.J., R.D., and P.R. contributed to the interpretation of findings. S.J. and P.R. drafted the manuscript. All authors provided critical feedback and substantially contributed to the revision of the manuscript. All authors read and approved the manuscript.

## Ethics declarations

## Acknowledgements

S.J. is supported by the 2023 Quad Fellowship. We acknowledge support in the form of computational credits from the Microsoft Accelerating Foundation Models Research (AFMR) grant.

## Extended Data

### Expert Sanity Checking of Agents

As part of the CRAFT-MD framework, dermatology experts annotated 120 GPT-4 and GPT-3.5 multi-turn conversations to sanity-check the behavior of the patient-AI and grader-AI agents, suggesting an opportunity for continued improvement of these agents.

#### Patient AI Reliability

An important part of realistic clinical conversations is to have patients responding in an accessible way using everyday language rather than complex medical terminology. We found that 13.3% of GPT-4 conversations and 10% of GPT-3.5 conversations were found to incorporate technical medical language in the patient-AI agent’s responses (Extended Data Figure 2). Examples include use of terms such as “pearly papules”, “diffuse shoddy lymphadenopathy”, and “seizure prophylaxis”. This still represents a lower level of technical medical language compared to the case vignettes, 100% of which contained specialized medical vocabulary. The patient AI agents appear to have partially learned to rephrase the vignette details into more layman-friendly responses when explicitly prompted. Furthermore, the patient-AI agent used phrases such as “the paragraph provided does not mention” in its responses (Figure 1), despite being prompted to not break character and reveal the use of a case vignette for describing symptoms. Finally, the patient-AI agent sometimes refused to provide requested information, offered only partial responses, or took over questioning instead of answering. Because the patient-AI agent was designed to strictly adhere to the case vignette information, the agent often responded with “I don’t have that information” to queries beyond the scope of the case vignette. While not prevalent, these examples of role confusion and selective unresponsiveness reveal gaps in comprehension and appropriate conversation dynamics.

#### Grader-AI Reliability

The grader-AI showed high correlation with dermatologist judgments on FRQ accuracy, as assessed by evaluating with three dermatology residents for different experimental setups. The average correlation was 0.94 for GPT-4 experiments and 0.912 for GPT-3.5 experiments (Supplementary Table 21; see Methods).

**Extended Data Figure 1:**
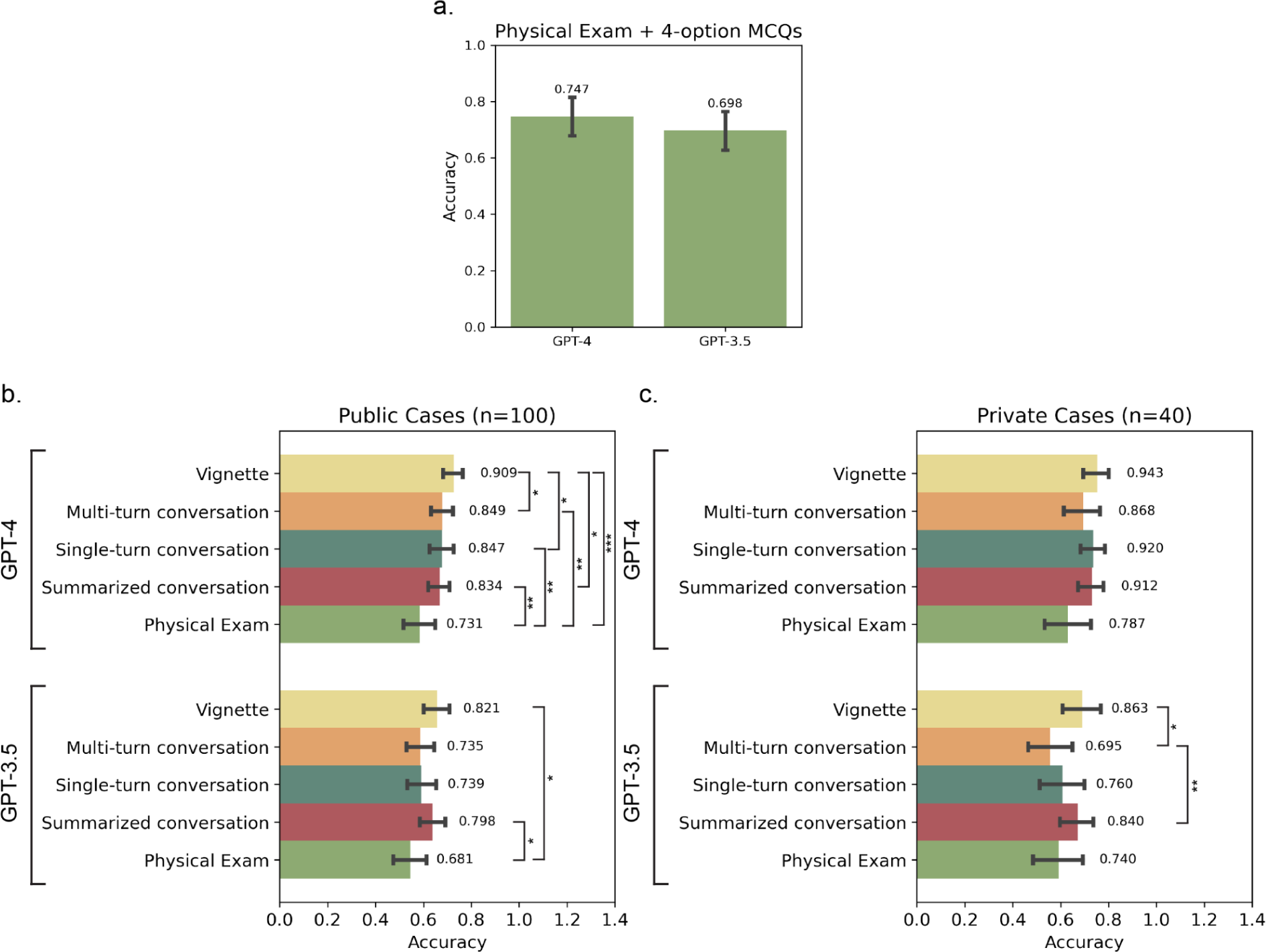
**(a)** Diagnostic accuracy for physical exam followed by 4-choice MCQs. **(b, c)** Diagnostic accuracy using GPT-4 and GPT-3.5, separated by public and private cases in the dataset, for five experimental setups: vignette + 4-choice MCQs, multi-turn conversation + 4-choice MCQs, single-turn conversation + 4-choice MCQs, summarized conversation + 4-choice MCQs, and physical exam + 4-choice MCQs. Error bars represent 95% confidence intervals, and numbers represent the mean accuracy. Statistically significant results have been annotated with brackets (* = <0.05, ** = <0.01, *** = <0.001).

**Extended Data Figure 2:**
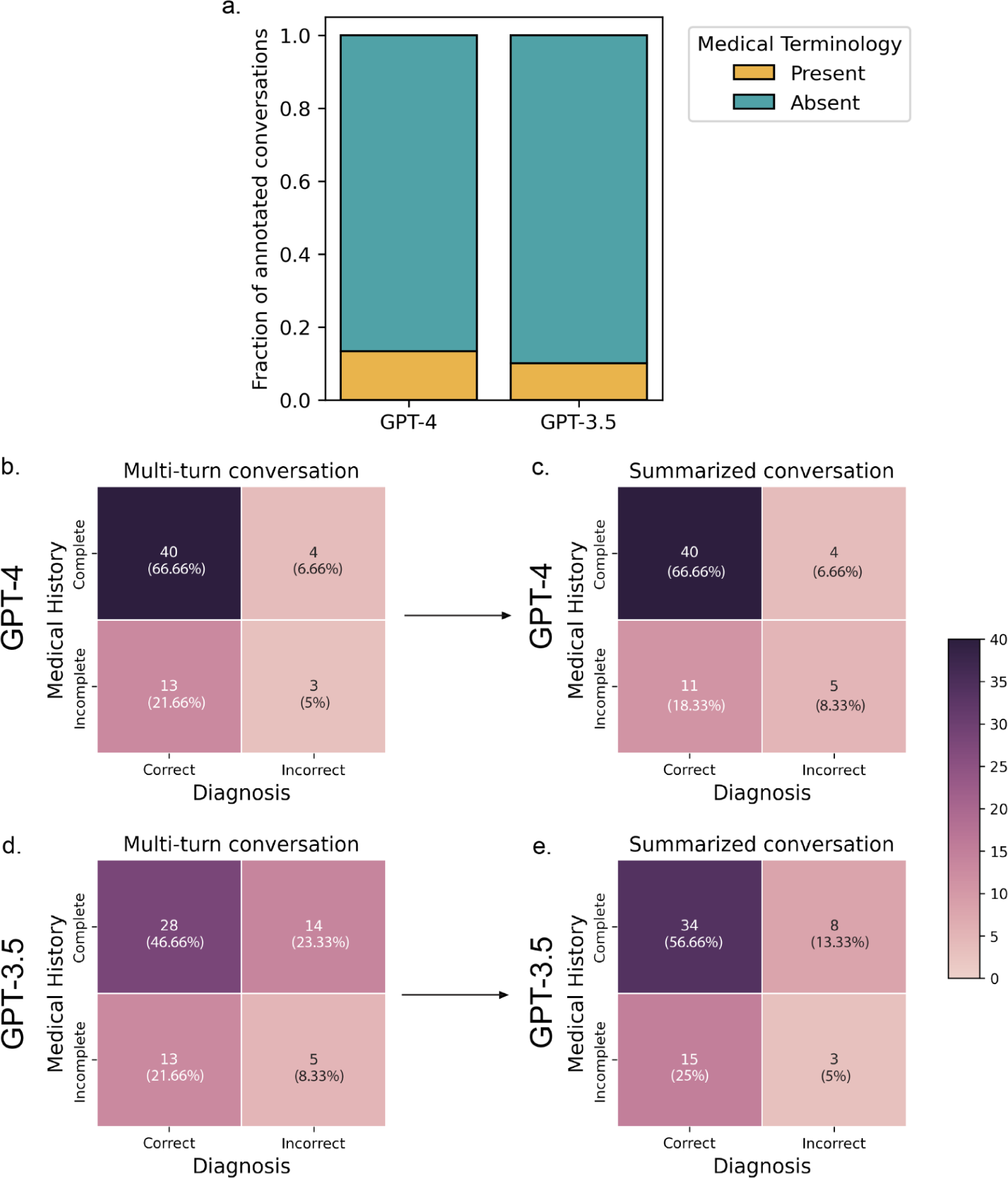
**(a)** Fraction of annotated GPT-4 and GPT-3.5 conversations with use of medical terminology by the patient-AI agent. Distribution of annotated GPT-4 **(b, c)** and GPT-3.5 **(d, e)** conversations for completeness of medical history extracted by the clinical LLM (evaluated by a senior dermatology resident) and the correctness of diagnosis (evaluated by grader-AI) in 4-choice MCQ setup.

**Extended Data Figure 3:**
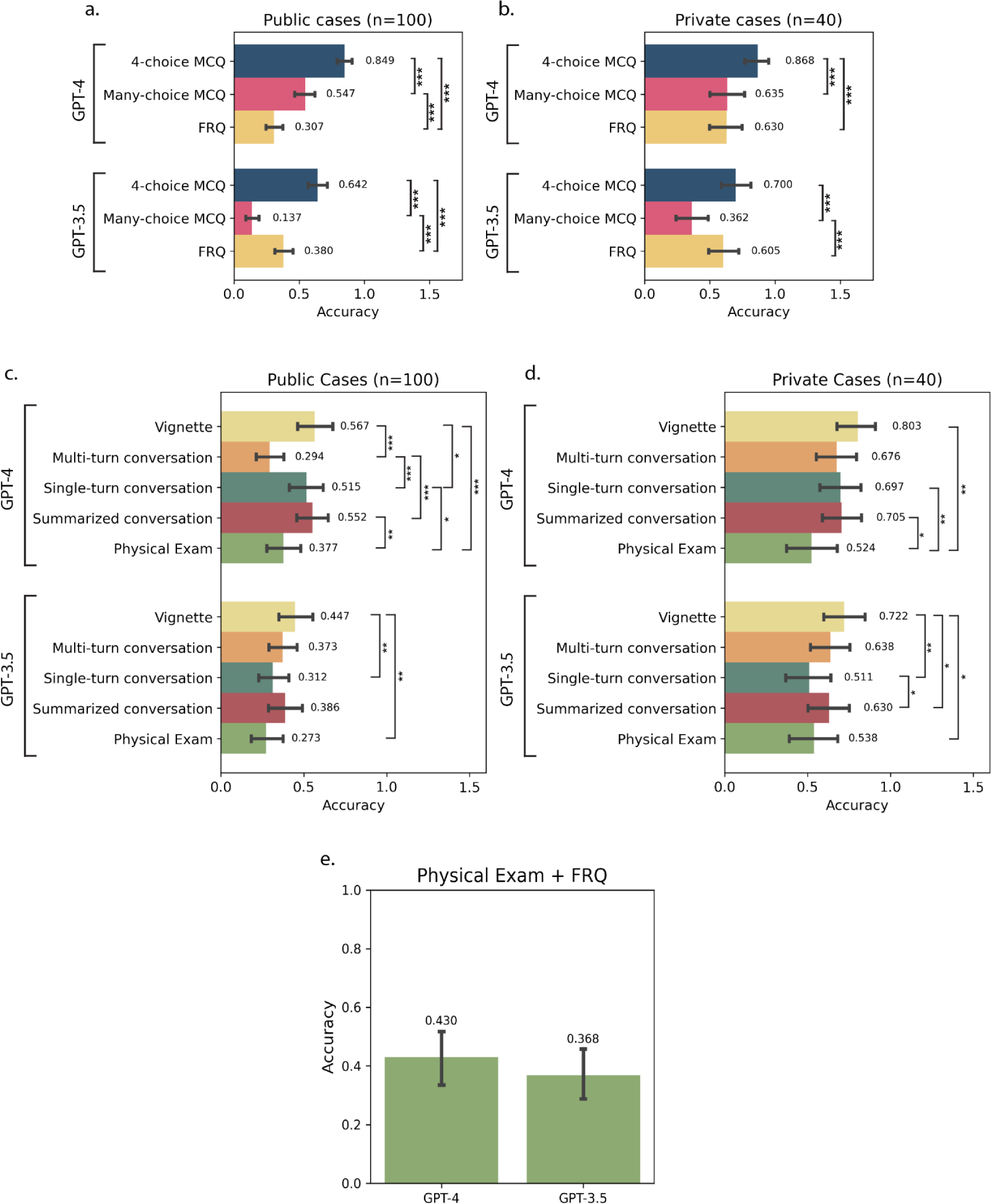
**(a, b)** Diagnostic accuracy using GPT-4 and GPT-3.5, separated by public and private cases in the dataset, when multi-turn conversation is followed by 4-choice, many-choice and no-choice MCQs. **(c, d)** Diagnostic accuracy using GPT-4 and GPT-3.5, separated by public and private cases in the dataset, for five experimental setups: vignette + FRQs, multi-turn conversation + FRQs, single-turn conversation + FRQs, summarized conversation + FRQs, and physical exam + FRQs. Error bars represent 95% confidence intervals, and numbers represent the mean accuracy. Statistically significant results have been annotated with brackets (* = <0.05, ** = <0.01, *** = <0.001). **(e)** Diagnostic accuracy for combined dataset using physical exam followed by FRQs.

**Extended Data Figure 4:**
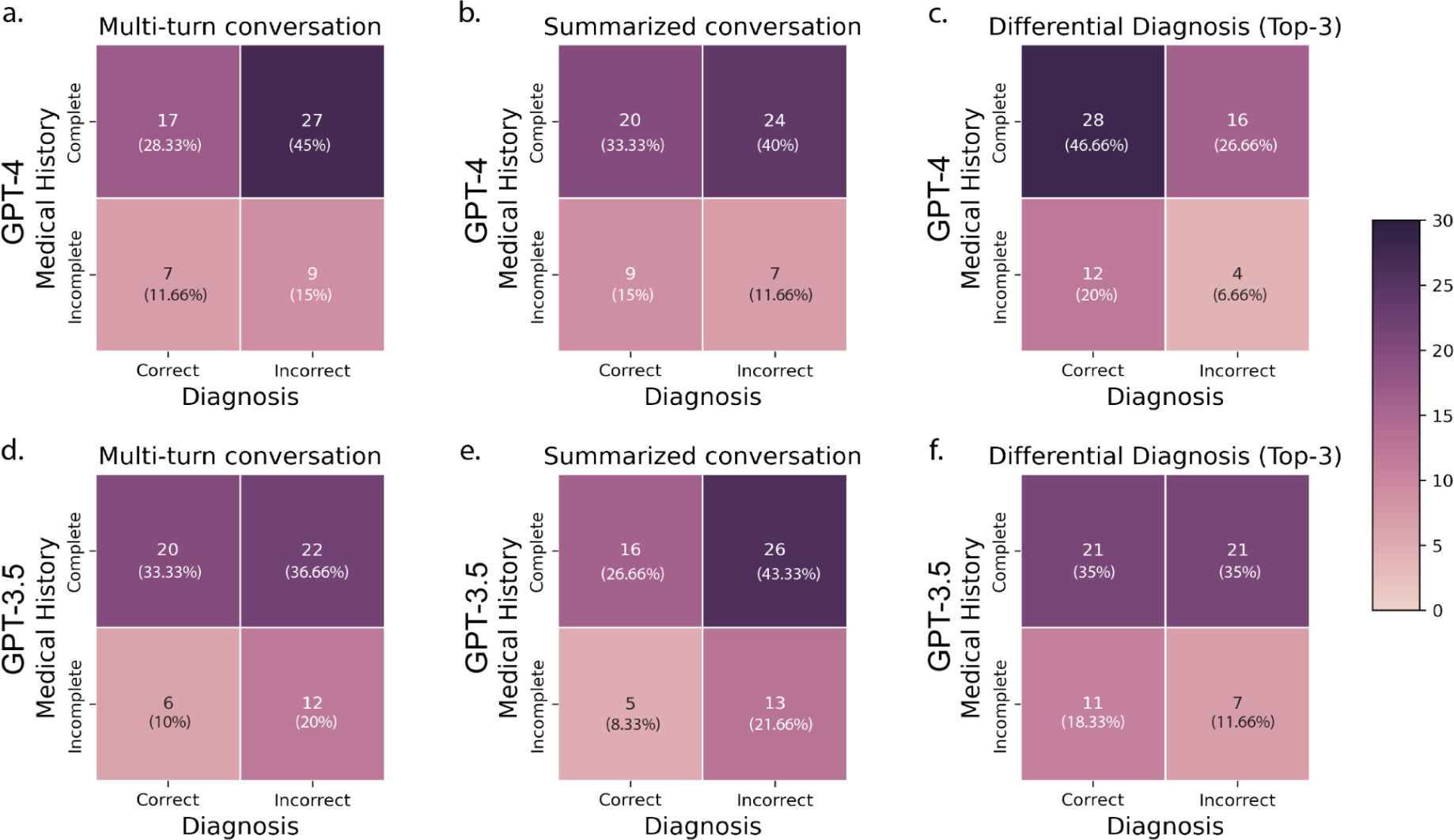
Distribution of annotated GPT-4 **(a, b, c)** and GPT-3.5 **(d, e, f)** conversations for completeness of medical history extracted by the clinical LLM (evaluated by a senior dermatology resident) and the correctness of diagnosis (evaluated by grader-AI) in **(a, d)** multi-turn conversation + no-choice MCQs. **(b, e)** summarized conversation + no-choice MCQs, and **(c, f)** multi-turn conversation + no-choice MCQs, when the clinical LLM is prompted to give the top-3 possible diagnoses instead of top-1.

## Supplementary Information

**Supplementary Table 1 :** Statistical significance for all cases (n=140) between different pairs of GPT-4 experiments for 4-choice MCQ.

| Experiment 1 | Experiment 2 | p-value | Adjusted p-value |
| --- | --- | --- | --- |
| Physical Exam | Multi-turn conversation | 2.00E-04 | 0.0018 |
| Physical Exam | Single-turn conversation | 2.00E-04 | 0.0018 |
| Physical Exam | Summarized conversation | 2.00E-04 | 0.0018 |
| Physical Exam | Vignette | 1.00E-04 | 0.001 |
| Multi-turn conversation | Single-turn conversation | 0.4303 | 1 |
| Multi-turn conversation | Summarized conversation | 0.8839 | 1 |
| Multi-turn conversation | Vignette | 0.0027 | 0.0135 |
| Single-turn conversation | Summarized conversation | 0.3676 | 1 |
| Single-turn conversation | Vignette | 0.0116 | 0.0464 |
| Summarized conversation | Vignette | 0.0018 | 0.0108 |

**Supplementary Table 2 :** Statistical significance for all public cases (n=100) between different pairs of GPT-4 experiments for 4-choice MCQ.

| Experiment 1 | Experiment 2 | p-value | Adjusted p-value |
| --- | --- | --- | --- |
| Physical Exam | Multi-turn conversation | 4.00E-04 | 0.0036 |
| Physical Exam | Single-turn conversation | 7.00E-04 | 0.0056 |
| Physical Exam | Summarized conversation | 7.00E-04 | 0.0056 |
| Physical Exam | Vignette | 1.00E-04 | 0.001 |
| Multi-turn conversation | Single-turn conversation | 0.9051 | 0.9051 |
| Multi-turn conversation | Summarized conversation | 0.2237 | 0.671 |
| Multi-turn conversation | Vignette | 0.0079 | 0.0325 |
| Single-turn conversation | Summarized conversation | 0.4016 | 0.8031 |
| Single-turn conversation | Vignette | 0.0065 | 0.0325 |
| Summarized conversation | Vignette | 0.0019 | 0.0114 |

**Supplementary Table 3 :** Statistical significance for all private cases (n=40) between different pairs of GPT-4 experiments for 4-choice MCQ.

| Experiment 1 | Experiment 2 | p-value | Adjusted p-value |
| --- | --- | --- | --- |
| Physical Exam | Multi-turn conversation | 6.07E-02 | 0.4249 |
| Physical Exam | Single-turn conversation | 5.50E-03 | 0.055 |
| Physical Exam | Summarized conversation | 6.30E-03 | 0.0567 |
| Physical Exam | Vignette | 1.61E-02 | 0.1288 |
| Multi-turn conversation | Single-turn conversation | 0.1595 | 0.7619 |
| Multi-turn conversation | Summarized conversation | 0.1524 | 0.7619 |
| Multi-turn conversation | Vignette | 0.1099 | 0.6593 |
| Single-turn conversation | Summarized conversation | 0.712 | 1 |
| Single-turn conversation | Vignette | 0.5994 | 1 |
| Summarized conversation | Vignette | 0.4213 | 1 |

**Supplementary Table 4 :** Statistical significance for all cases (n=140) between different pairs of GPT-3.5 experiments for 4-choice MCQ.

| Experiment 1 | Experiment 2 | p-value | Adjusted p-value |
| --- | --- | --- | --- |
| Physical Exam | Multi-turn conversation | 0.4108 | 0.8471 |
| Physical Exam | Single-turn conversation | 8.03E-02 | 0.3212 |
| Physical Exam | Summarized conversation | 0.0004 | 0.0032 |
| Physical Exam | Vignette | 7.00E-04 | 0.0049 |
| Multi-turn conversation | Single-turn conversation | 0.3824 | 0.8471 |
| Multi-turn conversation | Summarized conversation | 1.00E-04 | 0.001 |
| Multi-turn conversation | Vignette | 2.00E-04 | 0.0018 |
| Single-turn conversation | Summarized conversation | 0.0025 | 0.015 |
| Single-turn conversation | Vignette | 0.0029 | 0.015 |
| Summarized conversation | Vignette | 0.2824 | 0.8471 |

**Supplementary Table 5 :** Statistical significance for public cases (n=100) between different pairs of GPT-3.5 experiments for 4-choice MCQ.

| Experiment 1 | Experiment 2 | p-value | Adjusted p-value |
| --- | --- | --- | --- |
| Physical Exam | Multi-turn conversation | 0.1736 | 0.5207 |
| Physical Exam | Single-turn conversation | 8.33E-02 | 0.3332 |
| Physical Exam | Summarized conversation | 0.0028 | 0.0252 |
| Physical Exam | Vignette | 1.40E-03 | 0.014 |
| Multi-turn conversation | Single-turn conversation | 0.8929 | 0.8929 |
| Multi-turn conversation | Summarized conversation | 9.40E-03 | 0.0658 |
| Multi-turn conversation | Vignette | 7.50E-03 | 0.06 |
| Single-turn conversation | Summarized conversation | 0.0196 | 0.1176 |
| Single-turn conversation | Vignette | 0.0208 | 0.1176 |
| Summarized conversation | Vignette | 0.384 | 0.7679 |

**Supplementary Table 6 :** Statistical significance for private cases (n=40) between different pairs of GPT-3.5 experiments for 4-choice MCQ.

| Experiment 1 | Experiment 2 | p-value | Adjusted p-value |
| --- | --- | --- | --- |
| Physical Exam | Multi-turn conversation | 0.321 | 0.9629 |
| Physical Exam | Single-turn conversation | 6.52E-01 | 1 |
| Physical Exam | Summarized conversation | 0.0279 | 0.2232 |
| Physical Exam | Vignette | 3.08E-02 | 0.2232 |
| Multi-turn conversation | Single-turn conversation | 0.0843 | 0.3372 |
| Multi-turn conversation | Summarized conversation | 7.00E-04 | 0.007 |
| Multi-turn conversation | Vignette | 2.30E-03 | 0.0207 |
| Single-turn conversation | Summarized conversation | 0.0494 | 0.247 |
| Single-turn conversation | Vignette | 0.0363 | 0.2232 |
| Summarized conversation | Vignette | 0.514 | 1 |

**Supplementary Table 7 :**
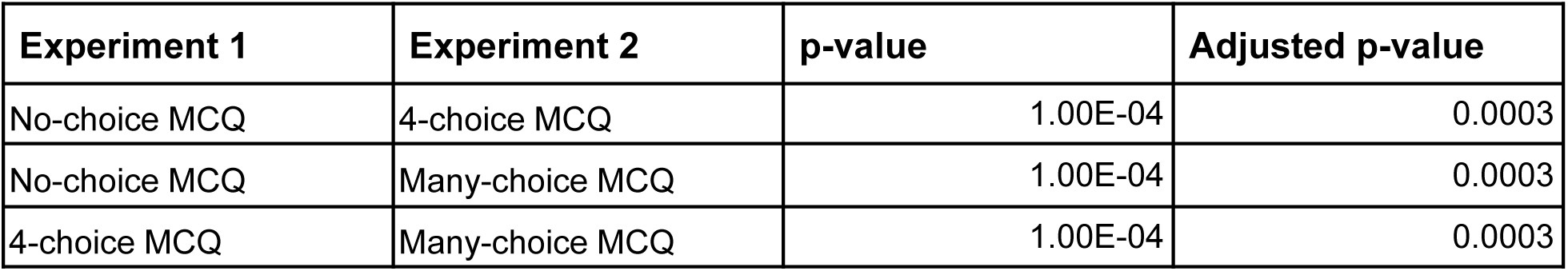
Statistical significance for all cases (n=140) between different pairs of GPT-4 experiments for multi-turn conversations.

**Supplementary Table 8 :**
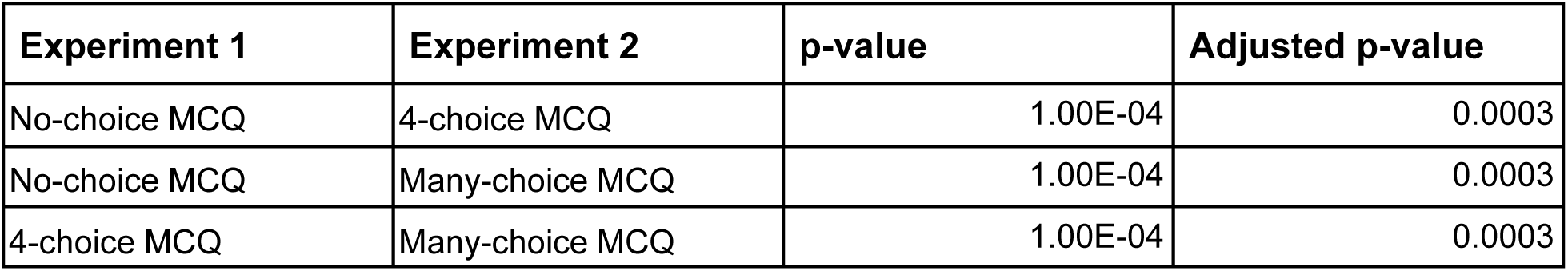
Statistical significance for public cases (n=100) between different pairs of GPT-4 experiments for multi-turn conversations.

**Supplementary Table 9 :**
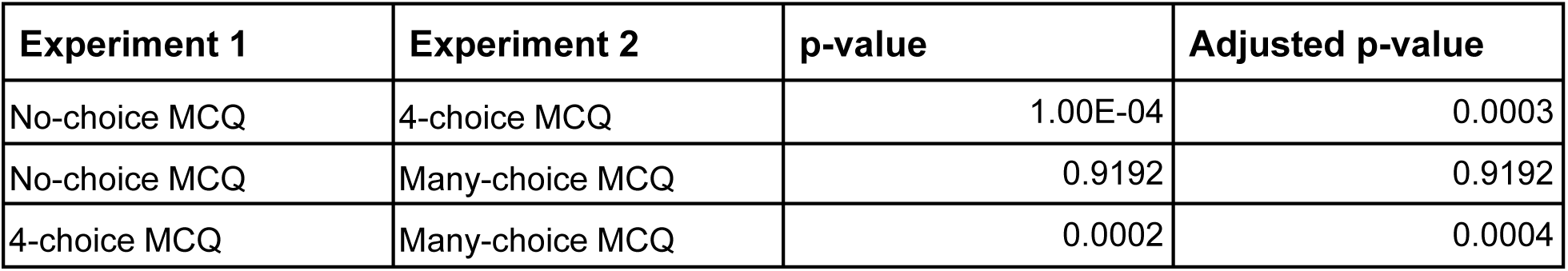
Statistical significance for private cases (n=40) between different pairs of GPT-4 experiments for multi-turn conversations.

**Supplementary Table 10 :**
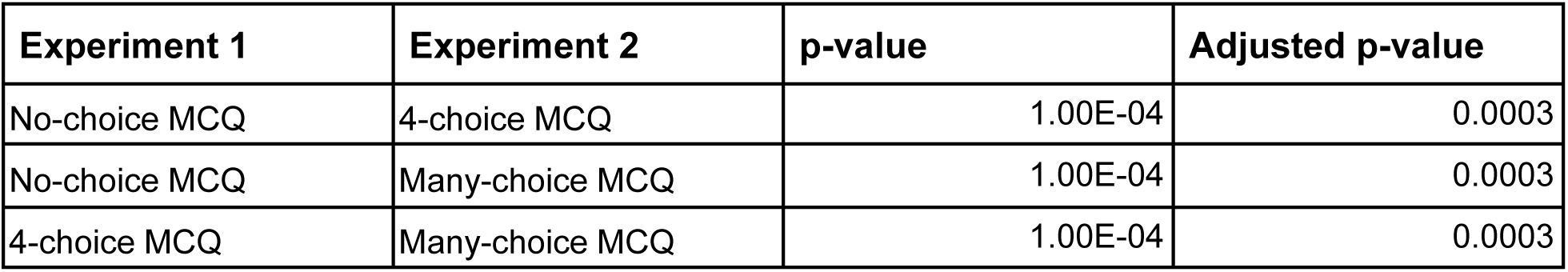
Statistical significance for all cases (n=140) between different pairs of GPT-3.5 experiments for multi-turn conversations.

**Supplementary Table 11 :**
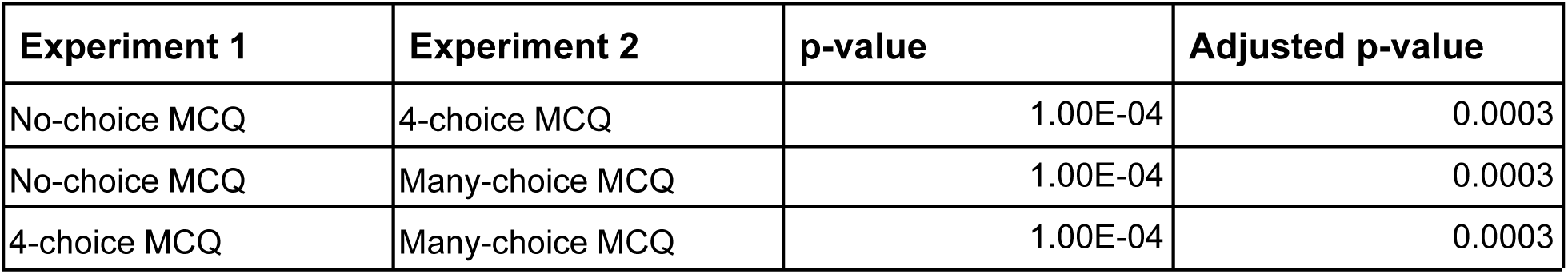
Statistical significance for public cases (n=100) between different pairs of GPT-3.5 experiments for multi-turn conversations.

**Supplementary Table 12 :** Statistical significance for private cases (n=40) between different pairs of GPT-3.5 experiments for multi-turn conversations.

| Experiment 1 | Experiment 2 | p-value | Adjusted p-value |
| --- | --- | --- | --- |
| No-choice MCQ | 4-choice MCQ | 0.1495 | 0.1495 |
| No-choice MCQ | Many-choice MCQ | 1.00E-04 | 0.0003 |
| 4-choice MCQ | Many-choice MCQ | 1.00E-04 | 0.0003 |

**Supplementary Table 13 :** Statistical significance for all cases (n=140) between different pairs of GPT-4 experiments for FRQs.

| Experiment 1 | Experiment 2 | p-value | Adjusted p-value |
| --- | --- | --- | --- |
| Physical Exam | Multi-turn conversation | 0.988 | 0.988 |
| Physical Exam | Single-turn conversation | 1.00E-04 | 0.001 |
| Physical Exam | Summarized conversation | 1.00E-04 | 0.001 |
| Physical Exam | Vignette | 1.00E-04 | 0.001 |
| Multi-turn conversation | Single-turn conversation | 1.00E-04 | 0.001 |
| Multi-turn conversation | Summarized conversation | 1.00E-04 | 0.001 |
| Multi-turn conversation | Vignette | 1.00E-04 | 0.001 |
| Single-turn conversation | Summarized conversation | 0.2369 | 0.4738 |
| Single-turn conversation | Vignette | 0.0003 | 0.0012 |
| Summarized conversation | Vignette | 0.004 | 0.012 |

**Supplementary Table 14 :** Statistical significance for public cases (n=100) between different pairs of GPT-4 experiments for FRQs.

| Experiment 1 | Experiment 2 | p-value | Adjusted p-value |
| --- | --- | --- | --- |
| Physical Exam | Multi-turn conversation | 0.201 | 0.402 |
| Physical Exam | Single-turn conversation | 2.20E-03 | 0.011 |
| Physical Exam | Summarized conversation | 2.00E-04 | 0.0012 |
| Physical Exam | Vignette | 1.00E-04 | 0.001 |
| Multi-turn conversation | Single-turn conversation | 1.00E-04 | 0.001 |
| Multi-turn conversation | Summarized conversation | 1.00E-04 | 0.001 |
| Multi-turn conversation | Vignette | 1.00E-04 | 0.001 |
| Single-turn conversation | Summarized conversation | 0.2194 | 0.402 |
| Single-turn conversation | Vignette | 0.0081 | 0.0324 |
| Summarized conversation | Vignette | 0.0561 | 0.1683 |

**Supplementary Table 15 :** Statistical significance for private cases (n=40) between different pairs of GPT-4 experiments for FRQs.

| Experiment 1 | Experiment 2 | p-value | Adjusted p-value |
| --- | --- | --- | --- |
| Physical Exam | Multi-turn conversation | 0.0153 | 0.0918 |
| Physical Exam | Single-turn conversation | 1.40E-03 | 0.0126 |
| Physical Exam | Summarized conversation | 3.30E-03 | 0.0264 |
| Physical Exam | Vignette | 3.00E-04 | 0.003 |
| Multi-turn conversation | Single-turn conversation | 5.04E-01 | 1 |
| Multi-turn conversation | Summarized conversation | 2.46E-01 | 0.7367 |
| Multi-turn conversation | Vignette | 8.20E-03 | 0.0574 |
| Single-turn conversation | Summarized conversation | 0.7834 | 1 |
| Single-turn conversation | Vignette | 0.0247 | 0.1225 |
| Summarized conversation | Vignette | 0.0245 | 0.1225 |

**Supplementary Table 16 :** Statistical significance for all cases (n=140) between different pairs of GPT-3.5 experiments for FRQs.

| Experiment 1 | Experiment 2 | p-value | Adjusted p-value |
| --- | --- | --- | --- |
| Physical Exam | Multi-turn conversation | 0.0061 | 0.0183 |
| Physical Exam | Single-turn conversation | 0.5138 | 1 |
| Physical Exam | Summarized conversation | 0.0007 | 0.0042 |
| Physical Exam | Vignette | 1.00E-04 | 0.001 |
| Multi-turn conversation | Single-turn conversation | 0.0036 | 0.018 |
| Multi-turn conversation | Summarized conversation | 0.8 | 1 |
| Multi-turn conversation | Vignette | 0.0045 | 0.018 |
| Single-turn conversation | Summarized conversation | 1.00E-04 | 0.001 |
| Single-turn conversation | Vignette | 1.00E-04 | 0.001 |
| Summarized conversation | Vignette | 1.00E-04 | 0.001 |

**Supplementary Table 17 :** Statistical significance for public cases (n=100) between different pairs of GPT-3.5 experiments for FRQs.

| Experiment 1 | Experiment 2 | p-value | Adjusted p-value |
| --- | --- | --- | --- |
| Physical Exam | Multi-turn conversation | 0.0346 | 0.173 |
| Physical Exam | Single-turn conversation | 0.1681 | 0.3362 |
| Physical Exam | Summarized conversation | 0.009 | 0.0588 |
| Physical Exam | Vignette | 7.00E-04 | 0.0063 |
| Multi-turn conversation | Single-turn conversation | 0.0941 | 0.2823 |
| Multi-turn conversation | Summarized conversation | 0.6193 | 0.6193 |
| Multi-turn conversation | Vignette | 0.0357 | 0.173 |
| Single-turn conversation | Summarized conversation | 6.60E-03 | 0.0528 |
| Single-turn conversation | Vignette | 4.00E-04 | 0.004 |
| Summarized conversation | Vignette | 8.40E-03 | 0.0588 |

**Supplementary Table 18 :** Statistical significance for private cases (n=40) between different pairs of GPT-3.5 experiments for FRQs.

| Experiment 1 | Experiment 2 | p-value | Adjusted p-value |
| --- | --- | --- | --- |
| Physical Exam | Multi-turn conversation | 0.0462 | 0.1848 |
| Physical Exam | Single-turn conversation | 0.5104 | 1 |
| Physical Exam | Summarized conversation | 0.0146 | 0.073 |
| Physical Exam | Vignette | 1.70E-03 | 0.0119 |
| Multi-turn conversation | Single-turn conversation | 0.0098 | 0.0588 |
| Multi-turn conversation | Summarized conversation | 0.8332 | 1 |
| Multi-turn conversation | Vignette | 0.0722 | 0.2166 |
| Single-turn conversation | Summarized conversation | 1.40E-03 | 0.0117 |
| Single-turn conversation | Vignette | 5.00E-04 | 0.005 |
| Summarized conversation | Vignette | 1.30E-03 | 0.0117 |

**Supplementary Table 19 :** Statistical significance for all cases (n=140) between different pairs of GPT-4 experiments for multi-turn conversations with and without physical exam (PE).

| Experiment 1 | Experiment 2 | p-value | Adjusted p-value |
| --- | --- | --- | --- |
| 4-choice MCQ | 4-choice MCQ (without PE) | 0.0002 | 0.0003 |
| many-choice MCQ | many-choice MCQ (without PE) | 1.00E-04 | 0.0003 |
| FRQ | FRQ (without PE) | 1.00E-04 | 0.0003 |

**Supplementary Table 20 :** Statistical significance for all cases (n=140) between different pairs of GPT-3.5 experiments for multi-turn conversations with and without physical exam (PE).

| Experiment 1 | Experiment 2 | p-value | Adjusted p-value |
| --- | --- | --- | --- |
| 4-choice MCQ | 4-choice MCQ (without PE) | 0.0006 | 0.0012 |
| many-choice MCQ | many-choice MCQ (without PE) | 0.3473 | 0.3473 |
| FRQ | FRQ (without PE) | 1.00E-04 | 0.0003 |

**Supplementary Table 21 :**
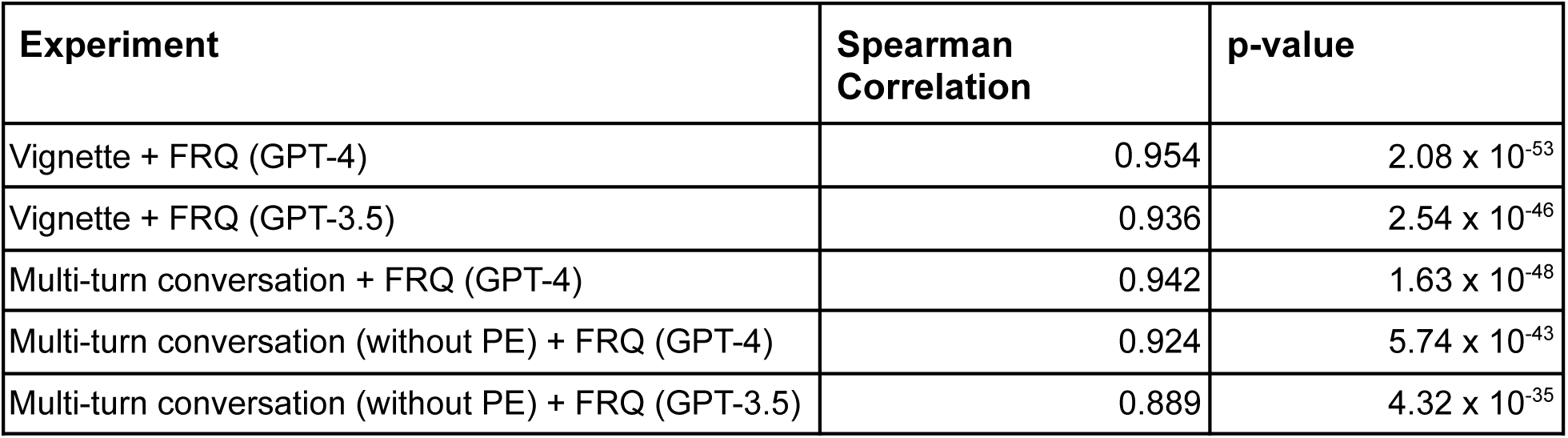
Correlation between grader-AI and dermatologists’ accuracy annotation of the clinical LLM (PE = physical exam, FRQ = Free Response Question).

